# Age-related Trust and its Association with Cost-related Medication Nonadherence

**DOI:** 10.64898/2026.11.09.26367639

**Authors:** James X. Zhang, David O. Meltzer

## Abstract

Trust operates foundationally at the personal and societal levels, a vital force that drives social connection, cooperation, and positive outcomes. In healthcare, trust is instrumental in improving patient outcomes as it forms the foundation for shared decision-making and patient-provider relationships. With growing evidence that generalized trust toward other people grows with age and behaviors such as cost-related medication non-adherence (CRN) decrease with age we study whether lack of trust is a force that drives up CRN and its variability in effects across age groups, controlling for other risk factors. The data consists of 680 Medicare diabetes patients at high risk of hospitalization enrolled in a randomized controlled trial of the effect of the Comprehensive Care Physician model (CCP) at an academic medical center between November 2012 and June 2016. This study provides first-of-its-kind evidence that lack of trust is an important force that drives up CRN and the effect of lack of trust decreased even though the lack of trust decreased by age increments. The effect of lack of trust is particularly pronounced among younger patients. For practice and policy, it shows the potential promising prospect that a more trusting attitude may be conducive to more productive health. More research is needed on how to build trust at the societal level and in healthcare specifically, and how generalized trust and target-specific trust of providers are correlated.

## Introduction

Trust operates foundationally at the personal and societal levels, a vital force that drives social connection, cooperation, and positive outcomes.^1,2^ In healthcare, trust is instrumental in improving patient outcomes as it forms the foundation for shared decision-making and patient-provider relationships.^3^ However, the mechanisms that underly the relationship between trust and outcomes are not well understood and warrant careful study.^4^ With growing evidence that generalized trust toward other people grows with age and behaviors such as cost-related medication non-adherence (CRN) decrease with age,^,5,6,7,8^ we study whether lack of trust is a force that drives up CRN and its variability in effects across age groups, controlling for other risk factors.

## Material and Methods

The data consists of 680 Medicare diabetes patients at high risk of hospitalization enrolled in a randomized controlled trial of the effect of the Comprehensive Care Physician model (CCP) at an academic medical center between November 2012 and June 2016. This was a period of relatively calm economic conditions between the Great Recession and the Covid pandemic without major policy shocks. This patient population was at increased risk for CRN, and the data included many known risk factors for CRN in addition to age for trust. The CCP study and data have been described elsewhere.^9,10^ The outcome measure is a 4-item questionnaire asking the respondents whether, in the past three months, they engaged in the following medication-related behaviors because the medicine cost too much: not filling or refilling a prescription, delaying getting a prescription filled, skipping doses to make the medicine last longer, and/or taking smaller doses to make the medicine last longer. An answer of “yes” to any of those 4 questions will be considered CRN. The measurement metric for trust is a generalized one based on the Structured Assessment of Personality Abbreviated Scale (SAPAS): it asks whether in general, the respondent trusts other people. We modeled a “no” answer to this question to show a clearer relationship between CRN and trust with age advancement in observed data and logistic regression models by controlling age and other covariates including gender, race, ethnicity, insurance status (Medicare-Medicaid dual eligible vs. non-dual), educational attainment, total household income before taxes (including salary, social security, pension, and other income), marital status, functional status (ADL & IADL deficiencies), chronic conditions (depression, cancer, stroke, kidney disease), institutionalization (NH residents), health literacy, and the CCP study arm.^9,10^ We first evaluated the association between trust and CRN controlling age and other covariates overall through a multivariable logit model with robust estimates; second, we used margins to calculate average marginal effects (AMEs) or interaction slopes between trust and CRN at each age group in the logit model.

## Results

A total of 163 (24%) patients reported CRN, and 224 (33%) answered “no” to the generalized trust questionnaire. The age groups are as follows: under 49 (101, 15%); 50-64 (192, 28%); 65-79 (310, 46%); 80+ (77, 11%). Other demographic variables are as follows: female (435, 64%), White (31, 5%); Black (606, 89%), other race (43, 6%), Hispanic (27, 4%), Medicare-Medicaid dual eligible (334, 49%), less than high school education (194, 29%). The income brackets are: <$2.5k (45, 7%); $2.5k-$5k (19, 3%); $5k-$10k (60, 9%); $10k-15k (154, 23%); $15k-$25k (74, 11%); $25k-$35k (31, 5%); $35k-$50k (46, 7%); D/K refusal (251, 37%). Figure 1 shows that both trust and CRN appeared to be age-related with downward trajectories with advancement from below 65 to 65+. Table 1 shows the adjusted odds ratio overall and average marginal effects (AMEs) of trust on CRN at each age group in the logit model controlling other covariates. A no-answer to the trust question was associated with a 62% increase in likelihood of CRN (p=0.03) compared to a yes-answer (Table 1). All AMEs were positive with statistical significance and showed a pattern of decreasing magnitude as age advanced.

**Table 1.**
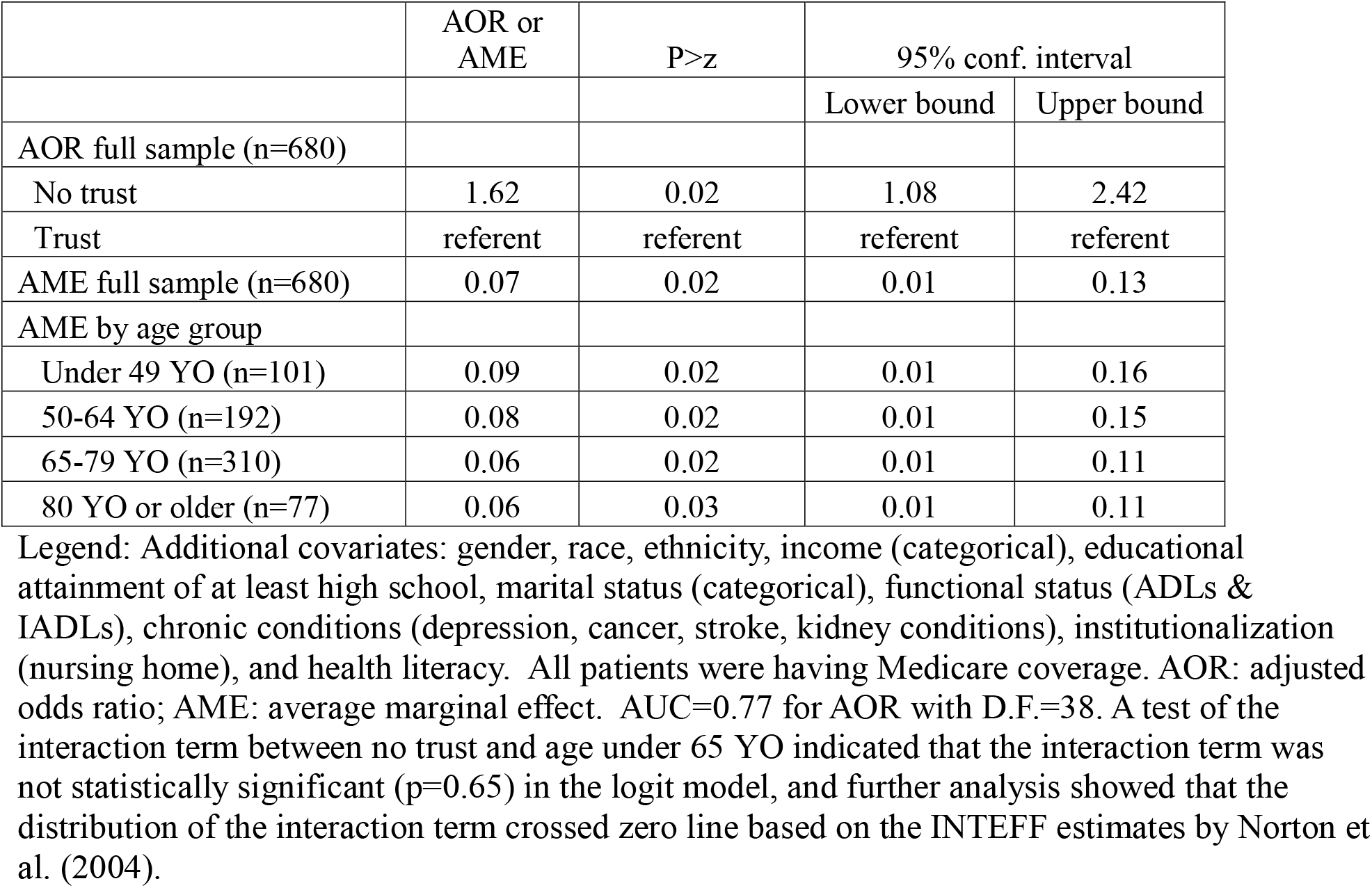
Adjusted Odds Ratio and Average Marginal Effect of No Trust on CRN.

|  | AOR or AME | P>z | 95% conf. interval |  |
|---|---|---|---|---|
|  |  |  | Lower bound | Upper bound |
| AOR full sample (n=680) |  |  |  |  |
| No trust | 1.62 | 0.02 | 1.08 | 2.42 |
| Trust | referent | referent | referent | referent |
| AME full sample (n=680) | 0.07 | 0.02 | 0.01 | 0.13 |
| AME by age group |  |  |  |  |
| Under 49 YO (n=101) | 0.09 | 0.02 | 0.01 | 0.16 |
| 50-64 YO (n=192) | 0.08 | 0.02 | 0.01 | 0.15 |
| 65-79 YO (n=310) | 0.06 | 0.02 | 0.01 | 0.11 |
| 80 YO or older (n=77) | 0.06 | 0.03 | 0.01 | 0.11 |

Legend: Additional covariates: gender, race, ethnicity, income (categorical), educational attainment of at least high school, marital status (categorical), functional status (ADLs & IADLs), chronic conditions (depression, cancer, stroke, kidney conditions), institutionalization (nursing home), and health literacy.  All patients were having Medicare coverage. AOR: adjusted odds ratio; AME: average marginal effect.  AUC=0.77 for AOR with D.F.=38. A test of the interaction term between no trust and age under 65 YO indicated that the interaction term was not statistically significant (p=0.65) in the logit model, and further analysis showed that the distribution of the interaction term crossed zero line based on the INTEFF estimates by Norton et al. (2004).

**Figure 1.**
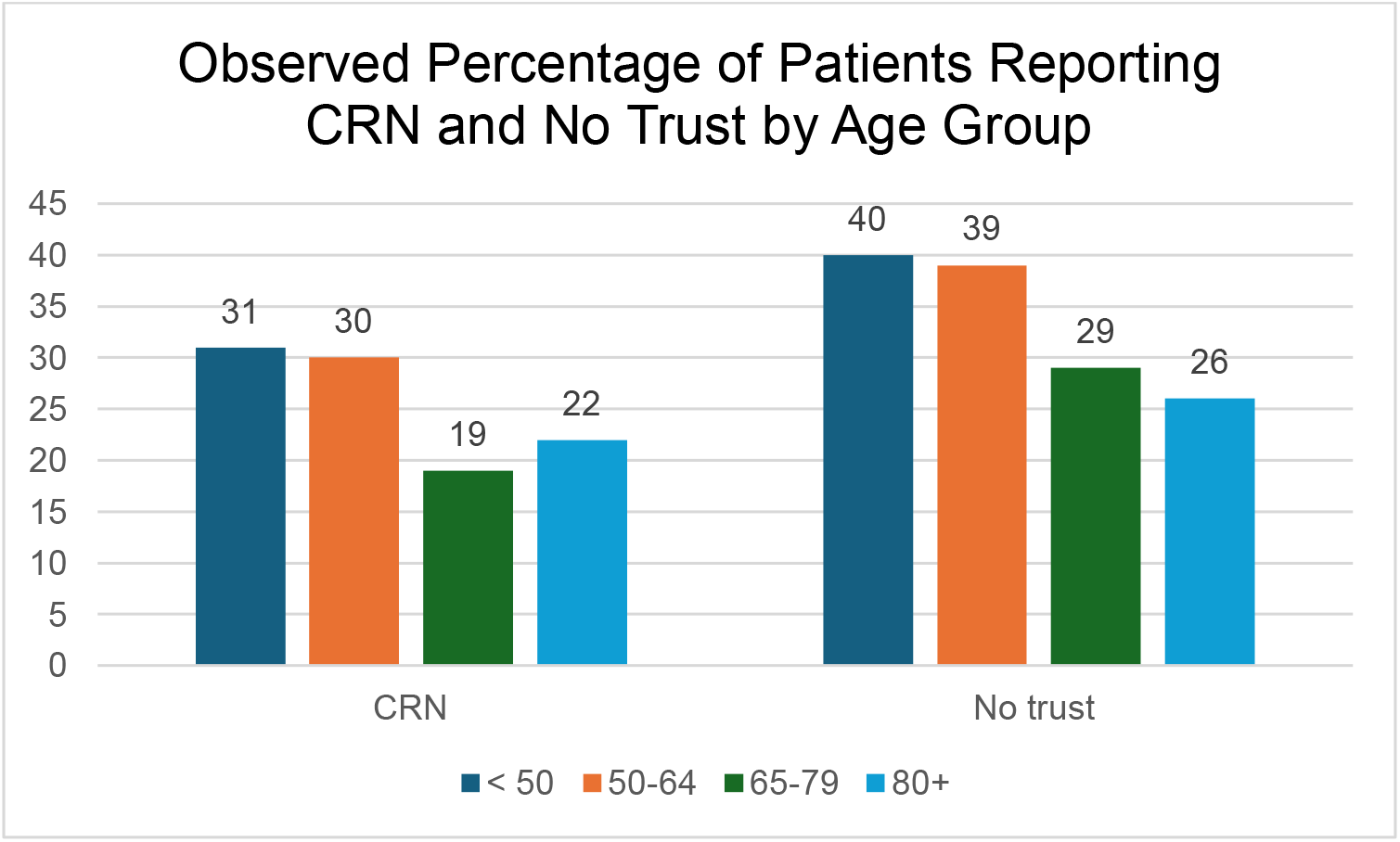
Observed Percentage of Patients Reporting CRN and No Trust by Age Group. Legend: P=0.01 and P=0.02 for CRN and No Trust by age group, respectively, by Chi-squared tests.

## Discussion and implications

This study provides first-of-its-kind evidence that lack of trust is an important force that drives up CRN and the effect of lack of trust decreased even though the lack of trust decreased by age increments. The effect of lack of trust is particularly pronounced among younger patients. The strength of the causal inference was further improved by using generalized trust instead of target-specific trust of providers as this captures the general force that influences both trust of providers and CRN. For practice and policy, it shows the potential promising prospect that a more trusting attitude may be conducive to more productive health. More research is needed on how to build trust at the societal level and in healthcare specifically, and how generalized trust and target-specific trust of providers are correlated. The study was limited by a sample of patients recruited in a single academic medical center. However, many known risk factors for CRN were controlled with a rich set of covariates to reduce potential confounding.

## Data Availability Statement

Deidentified, individual participant data underlying the text, tables, and figures of this article will be made available six months after article publication until five years after article publication. Researchers wishing to use these data must prepare a protocol for the use of these data which will be reviewed for methodological soundness and potential human subjects concerns by the study investigators and The University of Chicago Biological Sciences Division Institutional Review Board. Proposals should be directed to.

To gain access, data requestors will need to complete a data access agreement. Data and a data dictionary will be accessible through a secure website. Statistical code will be available six months after article publication until five years after article publication. Requests for code should be directed to.

